# Dental biofilm microbiota dysbiosis is associated with the risk of acute graft-versus-host disease after allogeneic hematopoietic stem cell transplantation

**DOI:** 10.1101/2021.02.04.21251019

**Authors:** Vitor Heidrich, Julia S. Bruno, Franciele H. Knebel, Vinícius C. de Molla, Wanessa Miranda-Silva, Paula F. Asprino, Luciana Tucunduva, Vanderson Rocha, Yana Novis, Celso Arrais-Rodrigues, Eduardo R. Fregnani, Anamaria A. Camargo

**Affiliations:** Centro de Oncologia Molecular, Hospital Sírio Libanês, São Paulo, SP, Brazil; Departamento de Bioquímica, Instituto de Química, Universidade de São Paulo, São Paulo, SP, Brazil; Centro de Oncologia, Hospital Sírio Libanês, São Paulo, SP, Brazil; Universidade Federal de São Paulo, São Paulo, SP, Brazil; Hospital das Clínicas da Faculdade de Medicina, Universidade de São Paulo/ICESP, São Paulo, SP, Brazil; Churchill Hospital, NHS-BT, Oxford, United Kingdom

## Abstract

Acute graft-versus-host disease (aGVHD) is one of the major causes of death after allogeneic hematopoietic stem cell transplantation (allo-HSCT). Recently, aGVHD onset was linked to intestinal microbiota (IM) dysbiosis. However, other bacterial-rich gastrointestinal sites, such as the mouth, which hosts several distinctive microbiotas, may also impact the risk of GVHD. The dental biofilm microbiota (DBM) is highly diverse and, like the IM, interacts with host cells and modulates immune homeostasis. We characterized changes in the DBM of patients during allo-HSCT and evaluated whether the DBM could be associated with the risk of aGVHD. DBM dysbiosis during allo-HSCT was marked by a gradual loss of bacterial diversity and changes in DBM genera composition, with commensal genera reductions and potentially pathogenic bacteria overgrowths. High *Streptococcus* and high *Corynebacterium* relative abundance at preconditioning were associated with a higher risk of aGVHD (67% vs. 33%; HR = 2.89, *P* = 0.04 and 73% vs. 37%; HR = 2.74, *P* = 0.04, respectively), while high *Veillonella* relative abundance was associated with a lower risk of aGVHD (27% vs. 73%; HR = 0.24, *P* < 0.01). *Enterococcus faecalis* bloom during allo-HSCT was observed in 20% of allo-HSCT recipients and was associated with a higher risk of aGVHD (100% vs. 40%; HR = 4.07, *P* < 0.001) and severe aGVHD (60% vs. 12%; HR = 6.82, *P* = 0.01). To the best of our knowledge, this is the first study demonstrating that DBM dysbiosis is associated with the aGVHD risk after allo-HSCT.

## INTRODUCTION

Allogeneic hematopoietic stem cell transplantation (allo-HSCT) is the only curative treatment for several hematologic diseases. However, allo-HSCT recipients may experience potentially fatal complications, such as infections and graft-versus-host disease (GVHD)^1^.

Acute GVHD (aGVHD) is a clinical syndrome characterized by maculopapular rash, hyperbilirubinemia, anorexia, diarrhea and abdominal pain^2^. The incidence of aGVHD grade II-IV is 30-40% at day 100^3^. During transplantation, chemotherapy, radiotherapy, and infection can damage host cells, releasing sterile damage-associated molecular patterns (DAMPs), and pathogen-associated molecular patterns (PAMPs) into the extracellular milieu. DAMPs and PAMPs activate donor T cells leading to a proinflammatory state. Simultaneously, donor regulatory T cells, myeloid-derived suppressor cells and tolerogenic dendritic cells are activated, counterbalancing the inflammation as an anti-inflammatory response. An imbalance in these events toward the proinflammatory state may result in aGVHD^4^.

In addition to graft source and the intensity of the conditioning regimen^4^, the intestinal microbiota (IM) composition was shown to be associated with the risk and intensity of aGVHD. Loss of IM diversity is observed during the pre- and post-transplantation period^5^, and low microbiota diversity at the time of stem cell engraftment has been associated with a higher risk of severe aGVHD^5^ and transplant-related death^6^.

Two non-exclusive ecological events can explain the link between loss of bacterial diversity and aGVHD risk: (1) absence or loss of protective commensal bacterial species and (2) sudden expansion (also known as bloom) of opportunistic pathogenic bacteria. Both events have been independently linked to aGVHD development. For instance, a higher abundance of commensal bacteria from the *Blautia* genus in the IM after allo-HSCT has been associated with reduced GVHD-related mortality and improved overall survival^7,8^. On the other hand, a shift in IM leading to the dominance of bacteria from the *Enterococcus* genus occurs more prominently in allo-HSCT recipients developing aGVHD^9^, and it is associated with increased GVHD-related mortality^10^.

Recent studies have shown that bacteria inhabiting the oral cavity can translocate to the gut^11^ and drive IM dysbiosis^12^. However, direct evaluation of the effect of allo-HSCT on the oral microbiota (OM) and the influence of OM dysbiosis on aGVHD risk have not been performed. To further understand the impact of gastrointestinal bacterial communities on aGVHD development following allo-HSCT, it would be crucial to extend the scope of these analyses to the OM.

The OM comprises over 700 bacterial species that stick to surfaces of the mouth, forming biofilms^13^. The dental biofilm microbiota (DBM), in particular, is among the richest and most diverse and, like the IM, interacts with host cells and modulates immune homeostasis^14^. In this study, we characterized changes of the DBM in patients during allo-HSCT and evaluated whether alterations in DBM diversity and composition could be associated with the risk of aGVHD.

## METHODS

### Sample collection and oral care protocol

Supragingival biofilm samples were collected from patients who underwent allo-HSCT. Samples were collected with sterile swabs at three phases during allo-HSCT: before the conditioning regimen (preconditioning), at aplasia and at engraftment. All patients were requested not to perform oral hygiene for at least 6h before sample collection. All patients were examined by an oral medicine specialist for potential infections and followed the same protocol for oral mucositis prophylaxis with photobiomodulation and oral hygiene with fluoride toothpaste and 0.12% chlorhexidine mouthwash. Informed consent was obtained from all participants prior to sample collection. The study was approved by the Institutional Ethics Committee (Protocol #1.414.217), in line with the Declaration of Helsinki.

### DNA extraction and sequencing

Bacterial cells were recovered from swabs by vortexing in TE buffer supplemented with PureLink RNAse A (Thermo Fisher Scientific, Waltham, MA, USA). DNA was extracted using the QIAamp DNA Blood Mini Kit (Qiagen, Hilden, Germany) according to the manufacturer’s protocol. Next, 12.5 ng of total DNA and pre-validated primers^15^ were used to amplify 16S rRNA hypervariable regions V3–V4. Amplicons were sequenced as described elsewhere^16^ on the MiSeq platform (Illumina, San Diego, CA, USA).

### Bioinformatics analyses

Reads were demultiplexed, and primer sequences were removed using the MiSeq Reporter software. Read processing was carried out within the QIIME 2 (*Quantitative Insights Into Microbial Ecology 2*) framework^17^. Briefly, forward and reverse sequences were filtered for quality and bimeras, denoised, and merged into consensus sequences with the DADA2 pipeline^18^, generating unique amplicon sequencing variants (ASVs). ASVs were further filtered for chimeric sequences using the SILVA database^19^ and UCHIME^20^, resulting in a total of 6 434 516 high-quality 16S rRNA sequences, with the median number of sequences obtained per sample being 58 867 (range: 2 153 - 240 734). Afterwards, ASVs were taxonomically assigned using the SILVA database and VSEARCH tool^21^.

### Microbiota and statistical analyses

As determined by per sample alpha diversity rarefaction curves, <12 500 read samples were considered defective and excluded. To adjust for differences in library sizes, the remaining samples were rarefied to 14 157 reads before calculating alpha diversity indexes (Shannon and Gini-Simpson indexes and the number of observed ASVs as a proxy for species richness) with the QIIME 2 *q2-diversity* plugin. Alpha diversity across transplantation phases was compared with the Mann-Whitney U test. The relative abundance of each genus was calculated with the QIIME 2 *q2-taxa* plugin. Differentially abundant genera across transplantation phases were identified using ANCOM^22^. ANCOM W represents the proportion of null hypotheses rejected when subtesting the differential abundance of a genus normalized by the abundance of each one of the genera in the dataset. W > 0.7 was considered as statistically significant. Cumulative incidence (CMI) rates for aGVHD (grade II to IV) and severe aGVHD (grade III and IV) were calculated with death as a competing event. Relative risks for developing aGVHD and severe aGVHD were estimated using the Fine-Gray risk regression model and adjusted for graft source and intensity of the conditioning regimen. Relative risks are presented as hazard ratios with 95% CIs and two-tailed P-values. R software (version 3.6.2) and the statistical package *cmprsk* (version 2.2.9) were used for statistical analyses.

## RESULTS

### Patient characteristics

A total of 30 patients who underwent allo-HSCT for hematologic disorders at Hospital Sírio Libanês between January 2016 and April 2018 were consecutively enrolled in our study. Patient clinical characteristics are summarized in Table 1. The most common underlying disease was acute leukemia (60%). The majority of patients received reduced-intensity conditioning (60%) and grafts from peripheral blood (67%).

**Table 1:** Clinical characteristics of study patients. HCT-CI, Hematopoietic cell transplantation-specific comorbidity index; MMF, mycophenolate mofetil; MTX, methotrexate; CsA, cyclosporin A; PTCy, post-transplant cyclophosphamide. *Acute leukemia: 11 acute myeloid leukemia and 7 acute lymphocytic leukemia cases; other: 5 non-Hodgkin lymphoma, 4 myelodysplastic syndrome, 1 chronic myeloid leukemia, 1 chronic lymphocytic leukemia and 1 multiple myeloma cases.

|  | **n = 30** |
| --- | --- |
| **Sex (Male)** | 16 (53%) |
| **Age in years (median, range)** | 50 (19-73) |
| **Underlying disease\*** |  |
| Acute leukemia | 18 (60%) |
| Other | 12 (40%) |
| **Conditioning intensity** |  |
| Reduced intensity | 18 (60%) |
| **Total body irradiation** | 11 (37%) |
| **Pre-transplant T-cell depletion** | 15 (50%) |
| **Graft source** |  |
| Bone marrow | 10 (33%) |
| Peripheral blood | 20 (67%) |
| **Donor** |  |
| Matched sibling | 9 (30%) |
| Haploidentical | 10 (33%) |
| Matched unrelated | 9 (30%) |
| Mismatched unrelated | 2 (7%) |
| **GVHD prophylaxis** |  |
| MMF + CsA | 11 (37%) |
| MTX + CsA | 10 (33%) |
| MMF + CsA + PTCy | 9 (30%) |
| **Follow- up in months (median, range)** | 37 (25-46) |

The standard antimicrobial prophylaxis in our institution included oral levofloxacin, antiviral prophylaxis with acyclovir or valacyclovir, and antifungal prophylaxis with echinocandins or azoles according to the patient’s risk of fungal infection. In addition, cephalosporin and antibiotic for anaerobic bacteria (metronidazole, meropenem or piperacillin/tazobactam) were administered to 70% and 57% of patients, respectively.

aGVHD was diagnosed and classified according to the Glucksberg grading system^23^. Fifteen patients developed grade II-IV aGVHD and, of those, 6 developed severe aGVHD (grade III-IV). None of this cohort’s clinical characteristics, including graft source, conditioning regimen, GVHD prophylaxis and antibiotics usage, were significantly associated with the risk of aGVHD [Table S1].

### Dental biofilm microbiota dysbiosis during allo-HSCT

Supragingival biofilm samples were collected for bacterial profiling at preconditioning, aplasia, and engraftment to characterize changes in DBM during allo-HSCT. Three engraftment samples were excluded from downstream analyses due to insufficient high-quality reads.

DBM alpha diversity was assessed using the Shannon index. We observed a statistically significant decrease in DBM alpha diversity during allo-HSCT [Figure 1A], with engraftment samples presenting the lowest overall bacterial diversity (median at each collection phase: 4.15, 3.39, and 2.75, respectively). A similar decrease in alpha diversity was observed when using the Gini-Simpson index [Figure S1A] or the number of observed ASVs as a proxy for species richness [Figure S1B].

**Figure 1:**
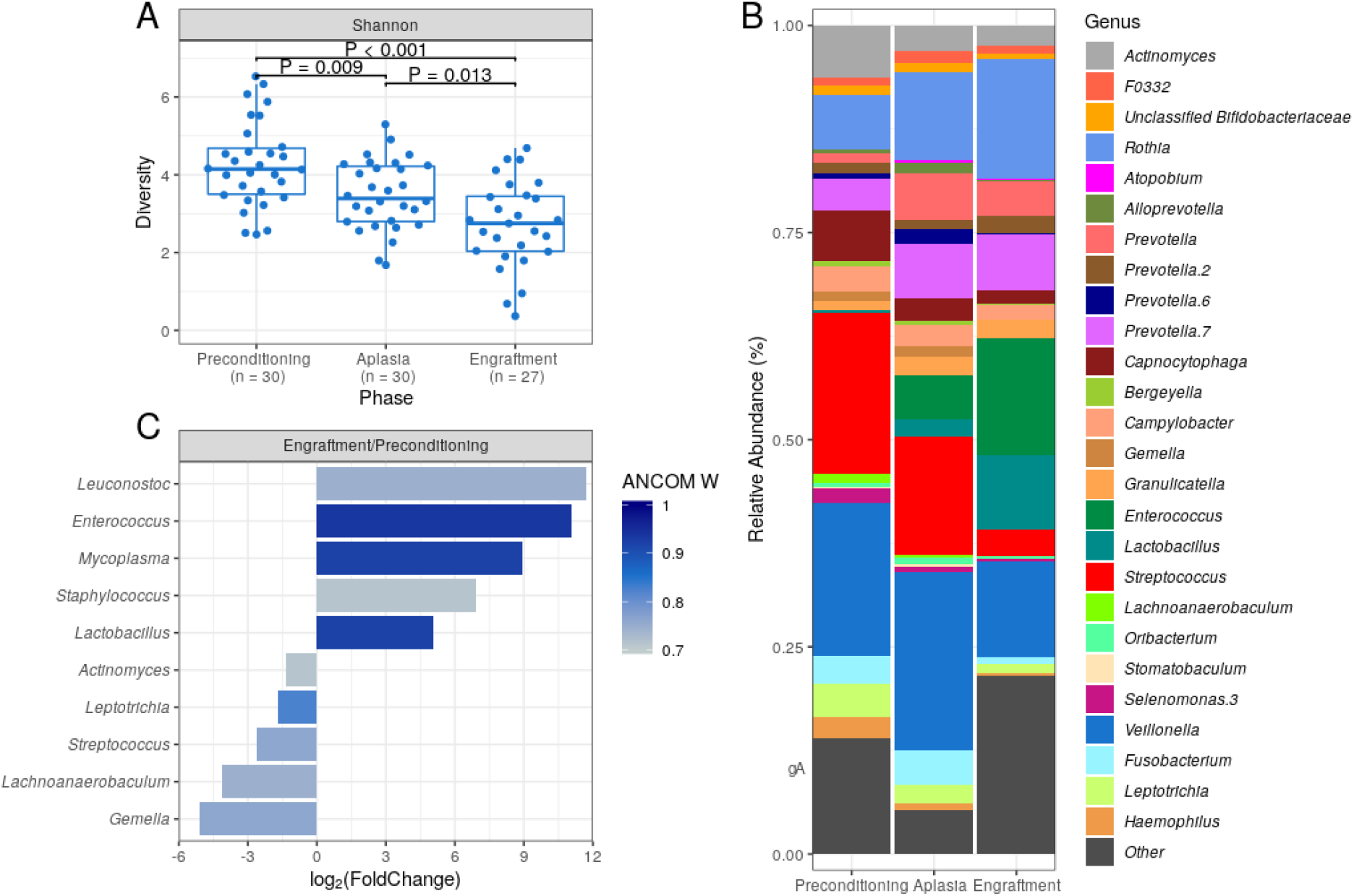
Characterization of dental biofilm microbiota (DBM) during allogeneic hematopoietic stem cell transplantation. (A) DBM alpha diversity (Shannon) boxplots at preconditioning (n = 30), aplasia (n = 30) and engraftment (n = 27). Mann-Whitney U test was used with the preconditioning as the reference for comparisons. The boxes highlight the median value and cover the 25th and 75th percentiles, with whiskers extending to the more extreme value within 1.5 times the length of the box. (B) Average DBM genera relative abundance composition across transplantation phases. Only genera with at least 0.1% relative abundance in at least 25% study samples are shown. Taxa are sorted based on taxonomic relatedness. (C) Significant genera relative abundance variations from preconditioning to engraftment according to ANCOM test (W > 0.7). Log_2_(Fold Change) for the average relative abundance variation (Engraftment/Preconditioning) is shown.

Marked changes in DBM genera composition were observed for all patients during allo-HSCT [Figure S2]. As expected, several dental biofilm commensal genera were detected at a high average relative abundance at preconditioning, including *Streptococcus* (19.5%), *Veillonella* (18.4%), *Actinomyces* (6.3%), and *Capnocytophaga* (6.1%) [Figure 1B]. However, their average relative abundance decreased during allo-HSCT. Likewise, we observed an increase in the average relative abundance of potentially pathogenic genera, such as *Enterococcus* and *Lactobacillus* [Figure 1B].

For a more quantitative assessment of DBM changes during allo-HSCT, we compared genera abundances at preconditioning and engraftment using the ANCOM test [Figure 1C]. The most statistically significant differences in abundance were observed for *Enterococcus, Lactobacillus*, and *Mycoplasma*, confirming the expansion of these potentially pathogenic genera in DBM during allo-HSCT. We also observed statistically significant (although less pronounced in terms of relative abundance change) decreases in commensal genera [Figure 1C].

### Dental biofilm microbiota diversity and aGVHD risk

Patients were stratified into two equal-sized groups (high and low-diversity groups) by the entire cohort’s median alpha diversity value to evaluate the association between DBM diversity and aGVHD risk. Using the Shannon diversity index, DBM diversity showed no association with the risk of aGVHD at preconditioning, aplasia, or engraftment [Figure 2A-C and Table 2]. Similar results were obtained when using the Gini-Simpson diversity index or the number of observed ASVs as a proxy for species richness [Figure S3].

**Table 2:** Univariate (non-adjusted) and adjusted competing risk analyses for the association of acute graft-versus-host disease with relevant microbiota variables. Each multivariate model adjusts for graft source and conditioning intensity. Statistically significant associations are emphasized. HR, Hazard ratio; CI, Confidence interval; P, preconditioning; A, aplasia; E, engraftment.

|  |  |  | Adjusted |  |  |  |  |  |  |  |  |  |
| --- | --- | --- | --- | --- | --- | --- | --- | --- | --- | --- | --- | --- |
|  | Non-adjusted |  | Veillonella at P |  | Streptococcus at P |  | Corynebacterium at P |  | Ratio at P |  | E.faecalis bloom |  |
|  | HR (95% CI) | P-value | HR (95% CI) | P-value | HR (95% CI) | P-value | HR (95% CI) | P-value | HR (95% CI) | P-value | HR (95% CI) | P-value |
| Graft source (Bone Marrow) | 0.95<br>(0.35-2.63) | 0.92 | 1.42<br>(0.43-9.03) | 0.38 | 0.75<br>(0.23-2.46) | 0.64 | 1.42<br>(0.40-5.04) | 0.59 | 0.78<br>(0.25-2.46) | 0.67 | 1.63<br>(0.42-6.35) | 0.49 |
| Conditioning intensity (Myeloablative) | 0.74<br>(0.26-2.17) | 0.59 | 0.50<br>(0.11-2.32) | 0.37 | 0.79<br>(0.24-2.61) | 0.7 | 0.79<br>(0.20-3.04) | 0.73 | 0.92<br>(0.27-3.16) | 0.89 | 0.94<br>(0.24-3.61) | 0.92 |
| Diversity (Shannon) at P (High vs. Low) | 0.68<br>(0.26-1.78) | 0.43 | - | - | - | - | - | - | - | - | - | - |
| Diversity (Shannon) at A (High vs. Low) | 0.88<br>(0.33-2.31) | 0.79 | - | - | - | - | - | - | - | - | - | - |
| Diversity (Shannon) at E (High vs. Low) | 0.92<br>(0.33-2.58) | 0.87 | - | - | - | - | - | - | - | - | - | - |
| **Veillonella at P** (High vs. Low) | 0.24<br>(0.08-0.70) | **0.009** | 0.21<br>(0.07-0.65) | **0.006** | - | - | - | - | - | - | - | - |
| **Streptococcus at P** (High vs. Low) | 2.89<br>(1.07-7.79) | **0.036** | - | - | 3.17<br>(1.12-9.01) | **0.03** | - | - | - | - | - | - |
| **Corynebacterium at P** (High vs. Low) | 2.74<br>(1.05-7.15) | **0.04** | - | - | - | - | 2.79<br>(0.99-7.9) | 0.053 | - | - | - | - |
| **Ratio at P** (>1 vs. ≤1) | 0.23<br>(0.08-0.62) | **0.004** | - | - | - | - | - | - | 0.22<br>(0.08-0.64) | **0.005** | - | - |
| Ratio at A (>1 vs. ≤1) | 0.45<br>(0.16-1.23) | 0.12 | - | - | - | - | - | - | - | - | - | - |
| Ratio at E (>1 vs. ≤1) | 0.73<br>(0.27-1.98) | 0.54 | - | - | - | - | - | - | - | - | - | - |
| Any genus bloom (Yes vs. No) | 2.29<br>(0.63-2.36) | 0.21 | - | - | - | - | - | - | - | - | - | - |
| **E. faecalis bloom** (Yes vs. No) | 4.07<br>(1.82-9.14) | **0.0007** | - | - | - | - | - | - | - | - | 4.90<br>(1.66-14.5) | **0.004** |

**Figure 2:**
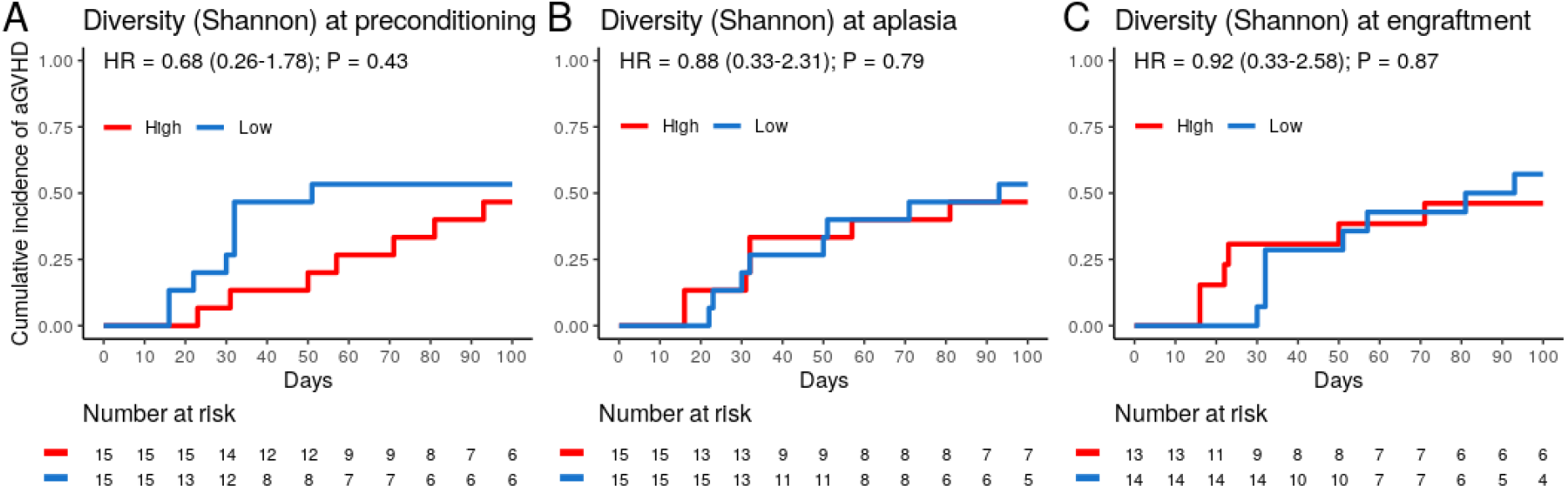
Dental biofilm microbiota alpha diversity is not associated with the risk of acute graft-versus-host disease (aGVHD). (A-C) Cumulative incidence of aGVHD with patients stratified by Shannon diversity index (High vs. Low) at preconditioning (A; n = 30), aplasia (B; n = 30) or engraftment (C; n = 27).

### Dental biofilm microbiota composition and aGVHD risk

We then evaluated whether the abundance of specific genera at preconditioning, aplasia, or engraftment was associated with the risk of aGVHD [Figure 3]. Only genera present at relative abundance ≥ 0.1% in at least 25% of the samples were considered for these analyses. Patients were stratified into two equal-sized groups (high and low relative abundance groups) by the median relative abundance observed in the entire cohort of each genus. *Veillonella, Streptococcus*, and *Corynebacterium* at preconditioning were significantly associated with the risk of aGVHD. We did not observe a similar association between the relative abundance of these or any other genus with the risk of aGVHD at aplasia or engraftment [Figure 3A].

**Figure 3:**
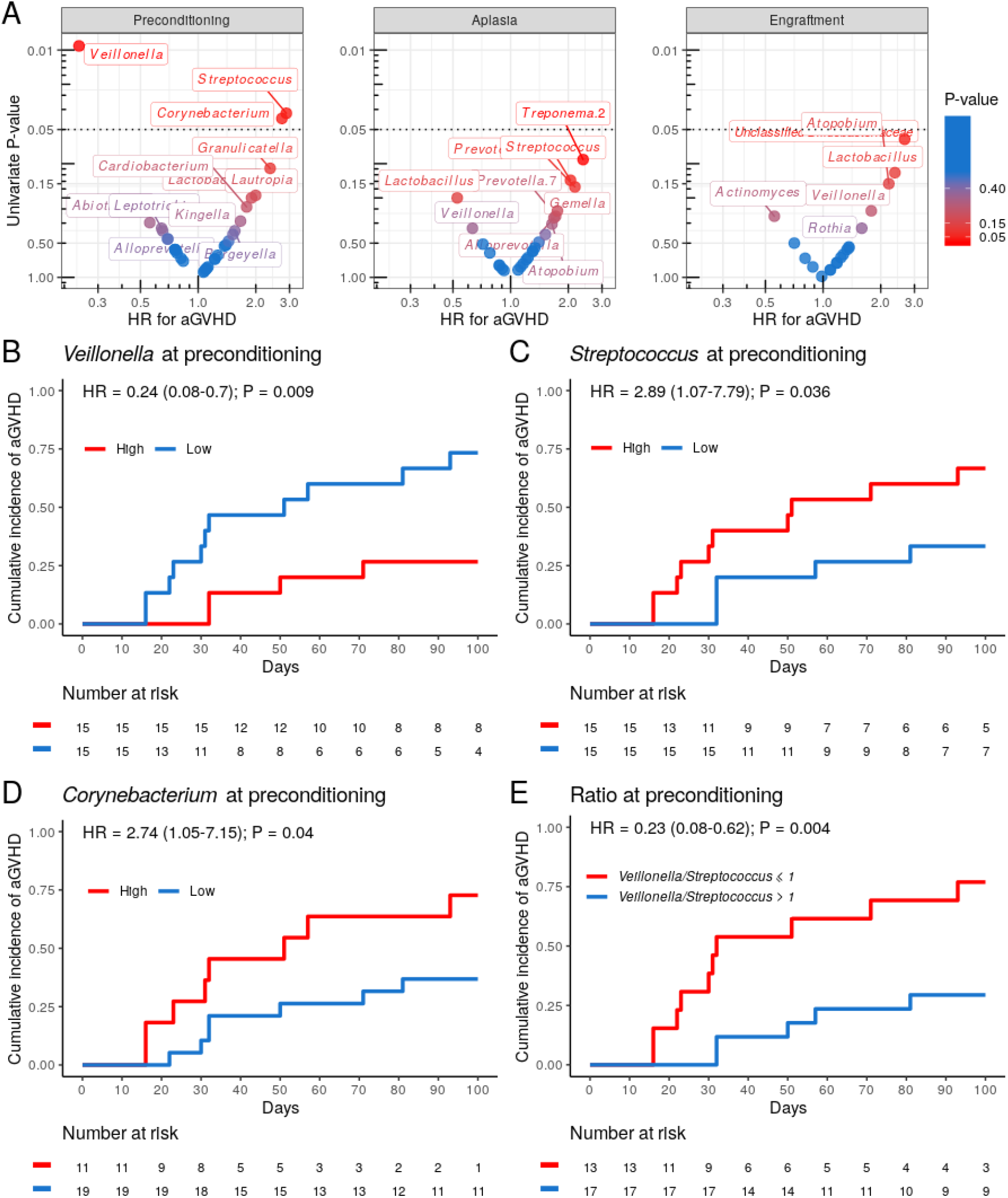
Specific genera relative abundance at preconditioning are associated with the risk of acute graft-versus-host disease (aGVHD). (A) Volcano plot for the univariate competing risk analysis for the association of aGVHD with genera relative abundance (hazard ratio vs. P-value) at preconditioning (left), aplasia (center) and engraftment (right). Only genera with ≥0.1% relative abundance in at least 25% of samples at a given phase were evaluated. Genera with P-value < 0.4 for the association are indicated explicitly. (B-D) Cumulative incidence of aGVHD with patients (n = 30) stratified by either *Veillonella* (B), *Streptococcus* (C) or *Corynebacterium* (D) relative abundance at preconditioning (High vs. Low). (E) Cumulative incidence of aGVHD with patients (n = 30) stratified by *Veillonella*/*Streptococcus* relative abundance ratio at preconditioning (>1 vs. ≤1).

Patients with high *Veillonella* relative abundance at preconditioning had a lower CMI of aGVHD (27% vs. 73%; HR = 0.24, 95% CI: 0.08–0.7, *P* = 0.009) [Figure 3B and Table 2]. This association remained significant after adjusting for graft source and intensity of the conditioning regimen (adjusted-HR = 0.21, 95% CI: 0.07–0.65, *P* = 0.006) [Table 2]. Patients with high *Streptococcus* or *Corynebacterium* relative abundance at preconditioning had a higher CMI of aGVHD (67% vs. 33%; HR = 2.89, 95% CI: 1.07–7.79, *P* = 0.036 and 73% vs. 37%; HR = 2.74, 95% CI: 1.05–7.15, *P* = 0.04, respectively) [Figure 3C and D; Table 2]. However, only *Streptococcus* remained significantly associated with risk of aGVHD after adjusting for graft source and intensity of the conditioning regimen (adjusted-HR = 3.17, 95% CI: 1.12–9.01, *P* = 0.03) [Table 2].

*Veillonella* and *Streptococcus* showed the highest average relative abundance at preconditioning [Figure 1B]. Given their overall high relative abundance and an inverse association with the risk of aGVHD, we next evaluated whether the *Veillonella*/*Streptococcus* ratio at preconditioning was associated with the risk of aGVHD. Patients with a *Veillonella*/*Streptococcus* ratio >1 at preconditioning had a lower CMI of aGVHD (29% vs. 77%; HR = 0.23, 95% CI: 0.08–0.62, *P* = 0.004) [Figure 3E and Table 2]. Interestingly, the association between the *Veillonella*/*Streptococcus* ratio at preconditioning and aGVHD risk was stronger than the association observed for each genus separately and remained significant after adjusting for graft source and intensity of the conditioning regimen (adjusted-HR = 0.22, 95% CI: 0.08–0.64, *P*=0.005) [Table 2]. The *Veillonella*/*Streptococcus* ratio at aplasia or engraftment was not associated with the risk of aGVHD [Table 2].

### Enterococcus faecalis bloom and aGVHD risk

Finally, we analyzed whether the blooming of potentially pathogenic genera observed during allo-HSCT was associated with the risk of aGVHD. For these analyses, bloom was defined as the sudden expansion of a particular genus from near absence (relative abundance <1% at preconditioning) to dominance (relative abundance ≥30% at aplasia or engraftment). Analyzing variations in genera relative abundance during allo-HSCT, we observed 23 blooms, involving 12 different genera and affecting a total of 20 patients. Three patients experienced more than one blooming event [Figure S4]. Patients experiencing any genus bloom (n = 20) did not have altered aGVHD risk [Table 2]. *Enterococcus* bloom was the most frequent event [Figure 4A], observed in 20% of the patients undergoing allo-HSCT. For all patients experiencing *Enterococcus* bloom except one, the phenomenon was attributed exclusively to *Enterococcus faecalis* expansion [Figure 4B]. There was no association between *E. faecalis* bloom and cephalosporin (Fisher exact test, *P* = 0.29) or antibiotic for anaerobic bacteria usage (Fisher exact test, *P* = 1).

**Figure 4:**
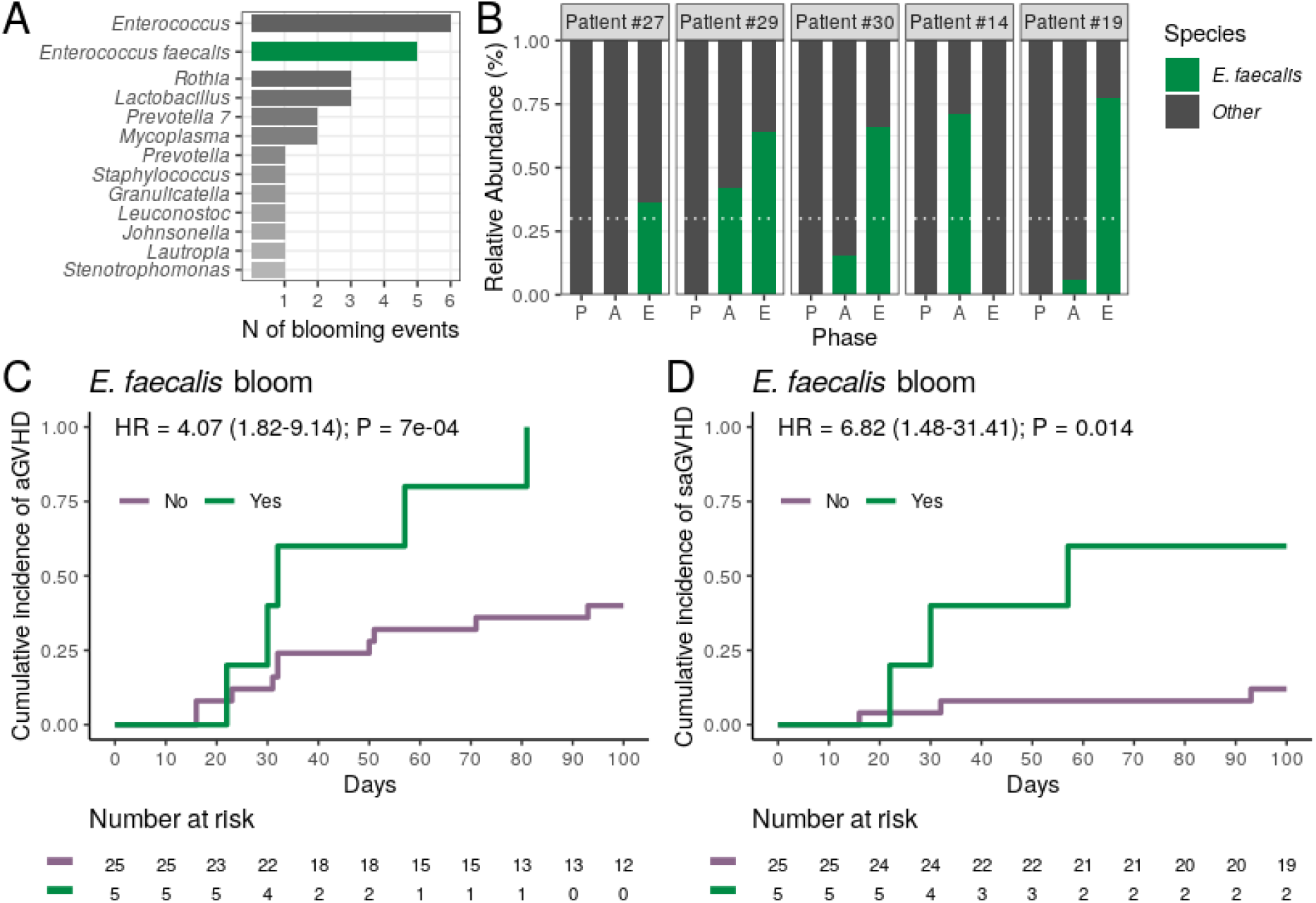
Dental biofilm *Enterococcus faecalis* bloom during allogeneic hematopoietic stem cell transplantation is associated with a higher risk of acute graft-versus-host disease (aGVHD) and severe aGVHD (saGVHD). (A) Number of observed blooming events per genera in all patients (n = 30). The number of *Enterococcus* blooms caused exclusively by *Enterococcus faecalis* is indicated. (B) Relative abundance of *Enterococcus faecalis* across transplantation phases for all patients experiencing *Enterococcus faecalis* bloom (n = 5). Patients are sorted based on the highest *Enterococcus faecalis* relative abundance observed per patient. White horizontal dashed line indicates dominance threshold. P, Preconditioning; A, Aplasia; E, Engraftment. (C-D) Cumulative incidence of aGVHD (C) or saGVHD (D) with patients (n = 30) stratified by *Enterococcus faecalis* bloom occurrence (No vs. Yes).

We next tested whether the occurrence of *E. faecalis* bloom was associated with the risk of aGVHD. All patients experiencing *E. faecalis* bloom developed aGVHD, and *E. faecalis* bloom was strongly associated with a higher CMI of aGVHD (100% vs. 40%; HR = 4.07, 95% CI: 1.82–9.14, *P* = 0.0007) [Figure 4C and Table 2]. This association remained significant after adjusting for graft source and intensity of the conditioning regimen (adjusted-HR = 4.90, 95% CI: 1.66–14.50, *P* = 0.004) [Table 2]. Notably, CMI of severe aGVHD (grade III-IV) was higher in patients experiencing *E. faecalis* bloom (60% vs. 12%; HR = 6.82, 95% CI: 1.48–31.41, *P* = 0.014) [Figure 4D and Table 2], revealing a direct association between DBM *E. faecalis* bloom and aGVHD risk and grade.

## DISCUSSION

In our study, we describe, for the first time using high-throughput 16S rRNA sequencing, changes in DBM diversity and composition in 30 patients undergoing allo-HSCT. As observed for IM, DBM dysbiosis during allo-HSCT was marked by a gradual loss of bacterial diversity, with engraftment samples presenting the lowest overall bacterial diversity. Like for IM, we also observed significant changes in DBM genera composition, with a decrease in the abundance of commensal core DBM genera, such as *Streptococcus* and *Actinomyces*^24^ (the only genera that can adhere to the tooth surface to start ordinary DB formation^24^), and overgrowths of potentially pathogenic bacteria, such as *Enterococcus, Lactobacillus*, and *Mycoplasma*. Most importantly, we observed that DBM genera relative abundance at preconditioning and changes in DBM composition during allo-HSCT (namely, *Enterococcus* bloom) were both predictive of aGVHD risk after allo-HSCT. There was no association between these aGVHD-associated microbiota variables and other allo-HSCT outcomes, including chronic GVHD [Table S2], as diagnosed in accordance with the NIH 2014 consensus^25^.

aGVHD is a major cause of non-relapse mortality following allo-HSCT, with a one-year survival rate for patients developing severe aGVHD of only 40%^26^. First-line therapy for aGVHD is based on corticosteroids, with response rates that vary between 40 and 70%^27^. In this scenario, identifying biomarkers capable of predicting aGVHD risk and developing preventive therapies is critical.

Recently, the IM composition has been analyzed as a biomarker for clinical outcomes in allo-HSCT recipients, including the development of aGVHD^5,8^. Moreover, microbiota-based therapeutic interventions, including microbiota-driven antibiotics selection, alternative dietary regimens (including probiotics/prebiotics usage) and fecal microbiota transplantation have been proposed to prevent and treat aGVHD^28–32^.

Like the IM, the OM plays an essential role in maintaining local and systemic health. Dental biofilm (DB) bacteria, as opposed to other shedding surface-living bacteria in the oral cavity, can adhere to hard surfaces and coaggregate^33^, allowing the assembly of an organized three-dimensional structure, which confers DBM its distinctive ecological properties. The DBM interacts directly with host immune cells and modulates immune homeostasis^14^. Moreover, DBM can also act as a microbial reservoir for systemic diseases. DBM dysbiosis can trigger local inflammation, destruction of surrounding periodontal tissue, and systemic translocation of oral microbes^24^. The influence of the OM in systemic diseases such as colorectal cancer^34^ and arthritis^35^ has been increasingly studied. However, in the allo-HSCT context, studies are still limited and have focused mainly on the saliva and the tongue microbiota^36–39^.

Loss of bacterial diversity in the salivary microbiota of patients undergoing allo-HSCT has been previously described and associated with oral mucositis^36^. Likewise, a steep decline in the tongue microbiota diversity was observed in severe aplastic anemia patients from preconditioning to the day of transplantation^37^. On the other hand, no appreciable changes in OM during allo-HSCT were observed in an additional study evaluating 4 different oral sites (buccal mucosa, saliva, tongue, and DB)^38^. However, this latter study used a low-resolution methodology (microarray) for microbiota characterization in a small number of patients (n = 11). Noteworthy, a single study evaluated the association between OM and allo-HSCT outcomes^39^. Allo-HSCT recipients showed a less diverse and distinct tongue microbiota on the day of transplantation than that of community-dwelling adults. In this study, the presence of the non-commensal bacteria *Staphylococcus haemolyticus* and/or *Ralstonia pickettii* in the tongue microbiota was significantly associated with lower overall survival after allo-HSCT, but not with aGVHD.

Out of the many allo-HSCT outcomes evaluated so far^40^, aGVHD onset has the clearest causal connection to the IM^28,29,40^. Briefly, it has been shown that the loss of SCFA-producing Clostridia species during the conditioning regimen reduces the concentration of intestinal butyrate^41^, a metabolite used as an energy source by intestinal epithelial cells^42^ and capable of promoting mucosal integrity^43^. Low butyrate levels promote extravasation of bacterial lipopolysaccharide through the damaged intestinal barrier, activating donor reactive T cells^40^. Additionally, lower butyrate levels correlate with lower abundance of regulatory T cells^44^ and impaired T-cell mediated local immune suppression^28^. Moreover, the loss of commensal bacteria that produce indole-3-aldehyde (including Clostridia class members), a metabolite that modulates barrier integrity by stimulating intestinal stem cell growth, is also linked to aGVHD development^45^. Finally, *Enterococcus faecalis* might contribute to aGVHD development via production of metalloproteases that impair barrier function^46^ and by stimulating macrophages to secrete TNF^47^. Accordingly, low IM diversity at the time of stem cell engraftment^6,8^, low abundance of commensal bacteria from Clostridia class members^7,8^, and intestinal enterococci dominance during allo-HSCT^10^ have been all associated with worsened aGVHD-related outcomes in studies evaluating stool specimens from allo-HSCT recipients^28,29,40^.

In our study, DBM diversity was not associated with the risk of aGVHD in any transplantation phase evaluated, which is in line with a recent IM study that did not find differences in IM diversity between aGVHD groups neither pre-nor post-transplantation^48^. Also, despite the presence (as expected^49^) of many Clostridia class genera in DBM (such as *Oribacterium*), we did not find DBM Clostridia class members significantly associated with the risk of aGVHD. However, as for IM, we observed a decrease in the relative abundance of several DB commensal genera during allo-HSCT, such as *Streptococcus, Veillonella, Actinomyces*, and *Capnocytophaga*, and an increase in the relative abundance of potentially pathogenic genera such as *Enterococcus* and *Lactobacillus*. Most importantly, high *Streptococcus* and high *Corynebacterium* relative abundance at preconditioning were associated with a higher risk of aGVHD, while high *Veillonella* relative abundance at preconditioning was associated with a lower risk of aGVHD.

Streptococci, corynebacteria, and veillonellae are part of the core DBM^50^ and represent the 1st, 2nd and 10th most important genera in terms of relative abundance in healthy volunteers DBM, respectively^49^. In our study, streptococci and veillonellae showed the highest average relative abundance at preconditioning and were both associated with the risk of aGVHD. Given their overall high relative abundance and the relative nature of the data, higher *Veillonella* relative abundance imposes lower *Streptococcus* relative abundance and vice versa. Hence, it is not possible to determine whether both genera are genuinely associated with the risk of aGVHD. Interestingly, the association between the *Veillonella/Streptococcus* ratio at preconditioning and aGVHD risk, independently of the conditioning regimen and graft source, was stronger than the association observed for each genus separately, suggesting a partial role for both genera in the observed effect.

During DB formation, bacterial early colonizers, after adhering to teeth salivary pellicles, coaggregate with other early and late colonizers, and a repeatable microbial succession takes place on the tooth surface^33^. Streptococci are the most abundant microbes in DB, representing a predominant early colonizer with broad coaggregation partnerships. Streptococci and veillonellae are in close physical contact during the early phases of DB maturation^33,51^ and can grow together in a metabolic cooperation-dependent manner^33,51^. Since this interaction occurs in the early phases of DB formation (and therefore are instrumental for DB maturation), the ratio *Veillonella/Streptococcus* might be a marker of early DBM disruption associated with a higher risk of aGVHD.

Corynebacteria bridge the early biofilm members to late colonizers^50^. In contradiction with the documented in the aforementioned healthy volunteers study^49^, we did not observe a high corynebacteria average relative abundance in any of the allo-HSCT phases evaluated. It is possible that the overall lower relative abundance of corynebacteria in detriment of early colonizers (such as streptococci and veillonellae) in our study may be indicative of a basal DBM disruption afflicting all allo-HSCT recipients. Alternatively, the lower relative abundance of corynebacteria may be explained by the stricter oral hygiene protocol recommended to our patients.

Finally, in our study, *E. faecalis* bloom in the DBM was observed in 20% of allo-HSCT recipients and was significantly associated with a higher risk of aGVHD and saGVHD. Noteworthy, despite recent in vitro evidence suggesting that high-dose of cephalosporin may promote *E. faecalis* biofilm formation^52^, there was no association between cephalosporin usage and DBM *E. faecalis* bloom in the evaluated cohort.

During allo-HSCT, intestinal enterococci expansion is well documented and is linked to both aGVHD development^10^ and subsequent bacteremia^53^. Notably, *E. faecalis* alone exacerbates aGVHD severity in gnotobiotic mouse models^10^. Our study reveals an additional site with enterococci expansion that might have systemic impacts after allo-HSCT. We can speculate that, during allo-HSCT, the dysbiotic DBM may act as an enterococci reservoir, triggering translocation to the gut and intestinal enterococci domination. This possibility is corroborated by the fact that there is intense oral bacteria translocation to the gut in hepatic cirrhosis patients^54^ and that such translocations in colorectal cancer patients are negatively correlated with intestinal Clostridia bacteria presence^34^. Indeed, oral bacteria translocation to the gut has been described in allo-HSCT recipients, and the presence of oral Actinobacteria and oral Firmicutes in stool samples of these patients was positively correlated with subsequent aGVHD development^5^. Alternatively, DBM enterococci may have an intestinal origin, since the injury to Goblet cells during conditioning regimen was shown to induce dissemination of dominant intestinal bacteria^28^. Further studies evaluating synchronously IM and DBM are necessary to decipher whether IM and DBM enterococci bloom are linked and which event precedes the other. Importantly, enterococci are present in small amounts in the healthy OM^49^ but may overgrow in pathogenic/dysbiotic settings, including after solid organ transplantation^55^, in a biofilm-dependent manner^56^. This may explain why previous microbiota studies on soft oral sites have not reported the expansion of *Enterococcus* in allo-HSCT recipients.

In conclusion, to our knowledge, this is the first study evaluating the DBM during allo-HSCT using a high-resolution technique. We identified markers of DBM dysbiosis during allo-HSCT. Most importantly, we showed that DBM composition during allo-HSCT may be predictive of aGVHD onset after transplantation, providing a simple and reproducible protocol for collection and analysis of allo-HSCT recipients microbiota before transplantation that may substitute fecal sampling when evaluating gastrointestinal dysbiosis and *Enterococcus* bloom. Nevertheless, our study has many limitations, especially being single-centered and having a limited sample size. Also, the study patients analyzed are heterogeneous and encompass several underlying diseases. Therefore, validation cohorts and prospective studies are needed to confirm our findings and test whether early interventions to correct DBM dysbiosis could prevent aGVHD onset. Further studies on DBM during allo-HSCT that include synchronous fecal sampling and metabolomics analyses are also needed to correlate our findings with aGVHD pathophysiology.

In conclusion, to our knowledge, this is the first study evaluating the DBM during allo-HSCT using a high-resolution technique. We identified markers of DBM dysbiosis during allo-HSCT. Most importantly, we showed that DBM composition during allo-HSCT may be predictive of aGVHD onset after transplantation, providing a simple and reproducible protocol for collection and analysis of allo-HSCT recipients microbiota before transplantation that may substitute fecal sampling when evaluating gastrointestinal dysbiosis and *Enterococcus* bloom. Nevertheless, our study has many limitations, especially being single-centered and having a limited sample size. Also, the study patients analyzed are heterogeneous and encompass several underlying diseases. Therefore, validation cohorts and prospective studies are needed to confirm our findings and test whether early interventions to correct DBM dysbiosis could prevent aGVHD onset. Further studies on DBM during allo-HSCT that include synchronous fecal sampling and metabolomics analyses are also needed to correlate our findings with aGVHD pathophysiology.

## Data Availability

Sequencing data will be publicly available as soon as the peer-reviewed version of the manuscript is published.

## Acknowledgements

V.H. was supported by Fundação de Amparo à Pesquisa do Estado de São Paulo (FAPESP, process no. 13996-0/2018). V.C.M. was supported by Conselho Nacional de Desenvolvimento Científico e Tecnológico (CNPq, process no. 141575/2018-2). J.S.B. was supported by Coordenação de Aperfeiçoamento de Pessoal de Nível Superior (CAPES, process no. 001).

## Notes

### Competing Interest Statement

The authors have declared no competing interest.

### Author Declarations

The study was approved by the ethics committee of Hospital Sírio-Libanês (approval number 1.414.217), in line with the Declaration of Helsinki.

### Summary of Updates

Criteria used for chronic graft-versus-host disease diagnosis and scoring included; better description of the underlying diseases in the study cohort provided (Table 1); references in the Discussion section updated.

