## Supplemental data for "Dental biofilm microbiota dysbiosis is associated with the risk of acute graft-versus-host disease after allogeneic hematopoietic stem cell transplantation"

### SUPPLEMENTARY INFORMATION
#### Supplementary tables

| | HR (95% CI) | P-value |
|---|---|---|
| Age (in years) | 0.99 (0.96-1.03) | 0.64 |
| Underlying disease* (AL vs. Other) | 1.00 (0.37-2.69) | 1 |
| Conditioning intensity (Myeloablative) | 0.74 (0.26-2.17) | 0.59 |
| TBI (Yes) | 0.79 (0.29-2.20) | 0.66 |
| T-cell depletion (Yes) | 0.78 (0.30-2.06) | 0.61 |
| Graft source (Bone Marrow) | 0.95 (0.35-2.63) | 0.92 |
| Donor (MSD vs. Haploidentical) | 1.14 (0.34-3.77) | 0.83 |
| Donor (MUD vs. Haploidentical) | 0.80 (0.24-2.73) | 0.72 |
| Donor (MMUD vs. Haploidentical) | 0.96 (0.14-6.30) | 0.96 |
| GVHD prophylaxis (MMF vs. MTX) | 1.32 (0.47-3.70) | 0.6 |
| GVHD prophylaxis (MMF+CyPT vs. MTX) | 1.18 (0.32-4.38) | 0.81 |
| AAB (Yes) | 0.92 (0.35-2.40) | 0.86 |
| Cephalosporin (Yes) | 3.44 (0.75-15.9) | 0.11 |

**Table S1**: **Univariate competing risk analysis for the association of acute graft-versus-host disease with clinical parameters**. All GVHD prophylaxis protocols include cyclosporin A. HCT-CI, Hematopoietic cell transplantation-specific comorbidity index; MMF, Mycophenolate mofetil; MTX, Methotrexate; TBI, Total body irradiation; AL, Acute leukemia; DRI, Disease relapse index; MSD, Matched sibling donor; MUD, Matched unrelated donor; MMUD, Mismatched unrelated donor; AAB, antibiotic for anaerobic bacteria; HR, Hazard ratio, CI, Confidence interval. *Acute leukemia: 11 acute myeloid leukemia and 7 acute lymphocytic leukemia cases**;** other: 5 non-Hodgkin lymphoma, 4 myelodysplastic syndrome, 1 chronic myeloid leukemia, 1 chronic lymphocytic leukemia and 1 multiple myeloma cases.

| | HR (95% CI) | P-value |
|---|---|---|
| Diversity (Shannon) at P (High vs. Low) | 0.89 (0.19-4.21) | 0.89 |
| Diversity (Shannon) at A (High vs. Low) | 0.18 (0.02-1.58) | 0.12 |
| Diversity (Shannon) at E (High vs. Low) | 0.92 (0.33-2.58) | 0.96 |
| *Veillonella* at P (High vs. Low) | 1.93 (0.35-10.6) | 0.45 |
| *Streptococcus* at P (High vs. Low) | 5.61 (0.67-47.1) | 0.11 |
| *Corynebacterium* at P (High vs. Low) | 0.95 (0.17-5.21) | 0.95 |
| Ratio at P (>1 vs. ≤1) | 0.68 (0.14-3.20) | 0.63 |
| Ratio at A (>1 vs. ≤1) | 1.12 (0.20-6.15) | 0.90 |
| Ratio at E (>1 vs. ≤1) | 0.60 (0.13-2.81) | 0.52 |
| Any genus bloom (Yes vs. No) | 0.97 (0.19-5.09) | 0.97 |
| *E. faecalis* bloom (Yes vs. No) | 1.02 (0.12-8.44) | 0.98 |

**Table S2**: **Univariate competing risk analysis for the association of chronic graft-versus-host disease with relevant microbiota variables**. HR, Hazard ratio; CI, Confidence interval; P, preconditioning; A, aplasia; E, engraftment.

#### Supplementary figures

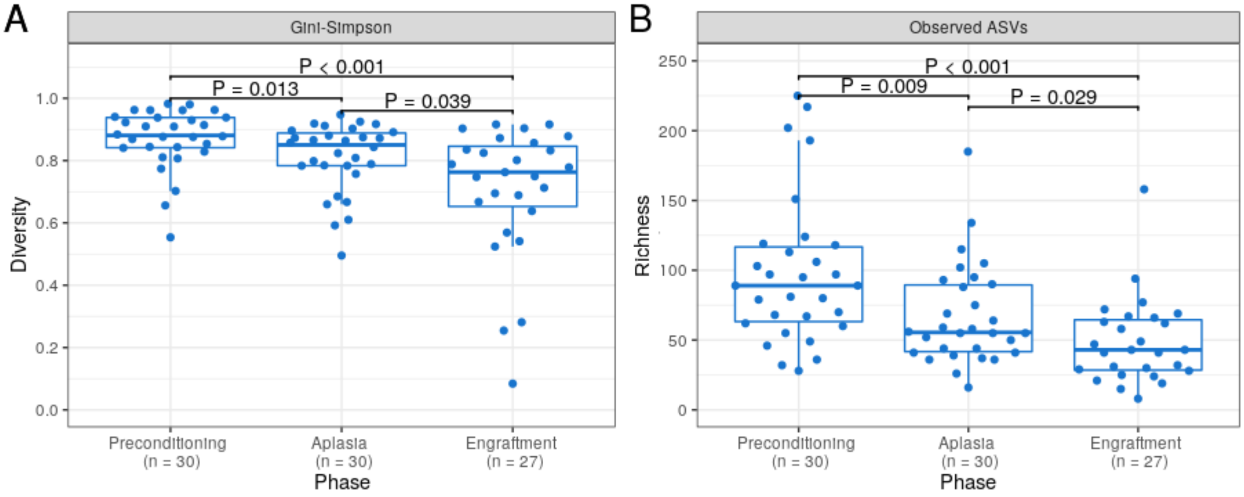


**Figure S1: Dental biofilm microbiota (DBM) alpha diversity decreases during allogeneic hematopoietic stem cell transplantation.** (A-B) DBM alpha diversity boxplots at preconditioning (n = 30), aplasia (n = 30) and engraftment (n = 27) as measured by either Gini-Simpson index (A) or the number of observed ASVs as a proxy for species richness (B). Mann-Whitney U test was used with the preconditioning as the reference for comparisons. The boxes highlight the median value and cover the 25th and 75th percentiles, with whiskers extending to the more extreme value within 1.5 times the length of the box.

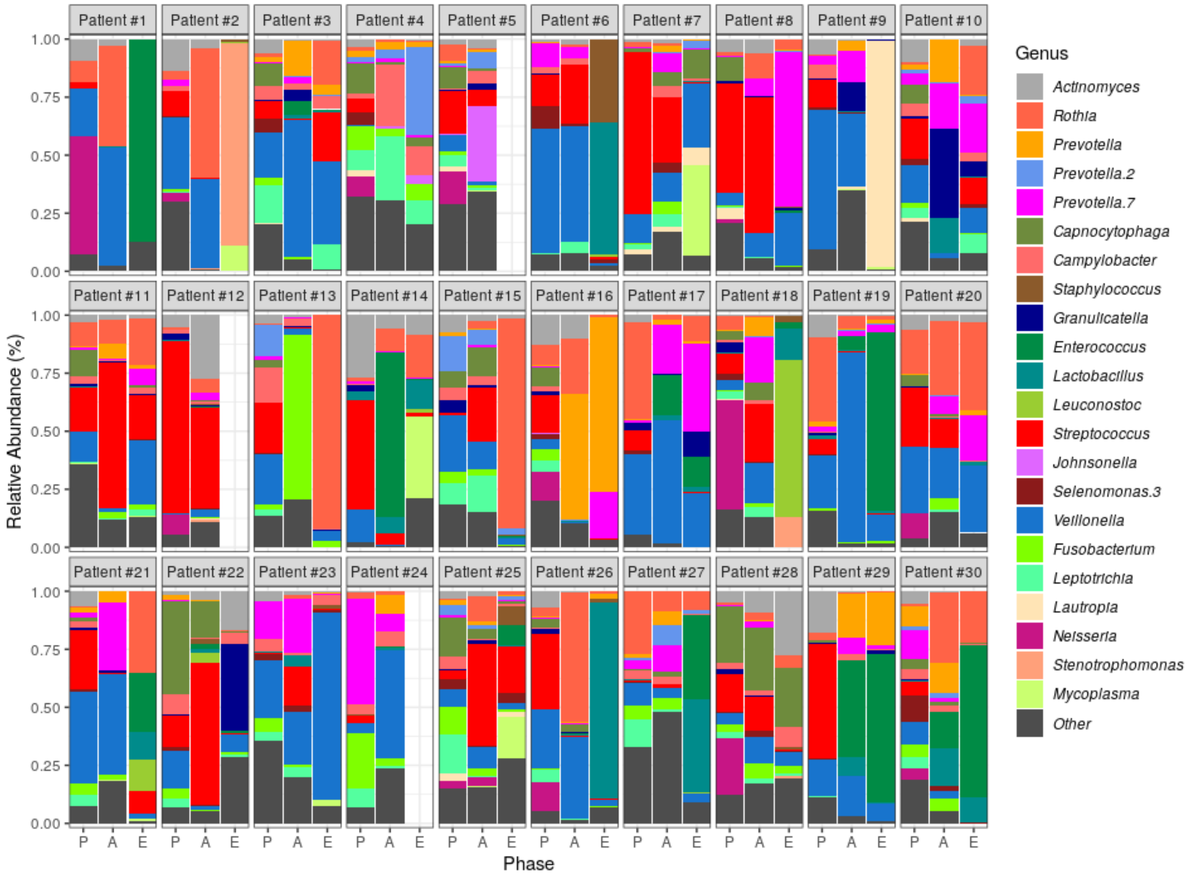


**Figure S2: Bacterial genera relative abundance changes in dental biofilm microbiota during allogeneic hematopoietic stem cell transplantation**. Genera relative abundance composition across transplantation phases for all patients (n = 30). Missing samples did not reach quality criteria for analyses. Only genera with at least 1% relative abundance in at least 25% study samples or dominant genera are shown. Taxa are sorted based on taxonomic relatedness. P, Preconditioning; A, Aplasia; E, Engraftment.

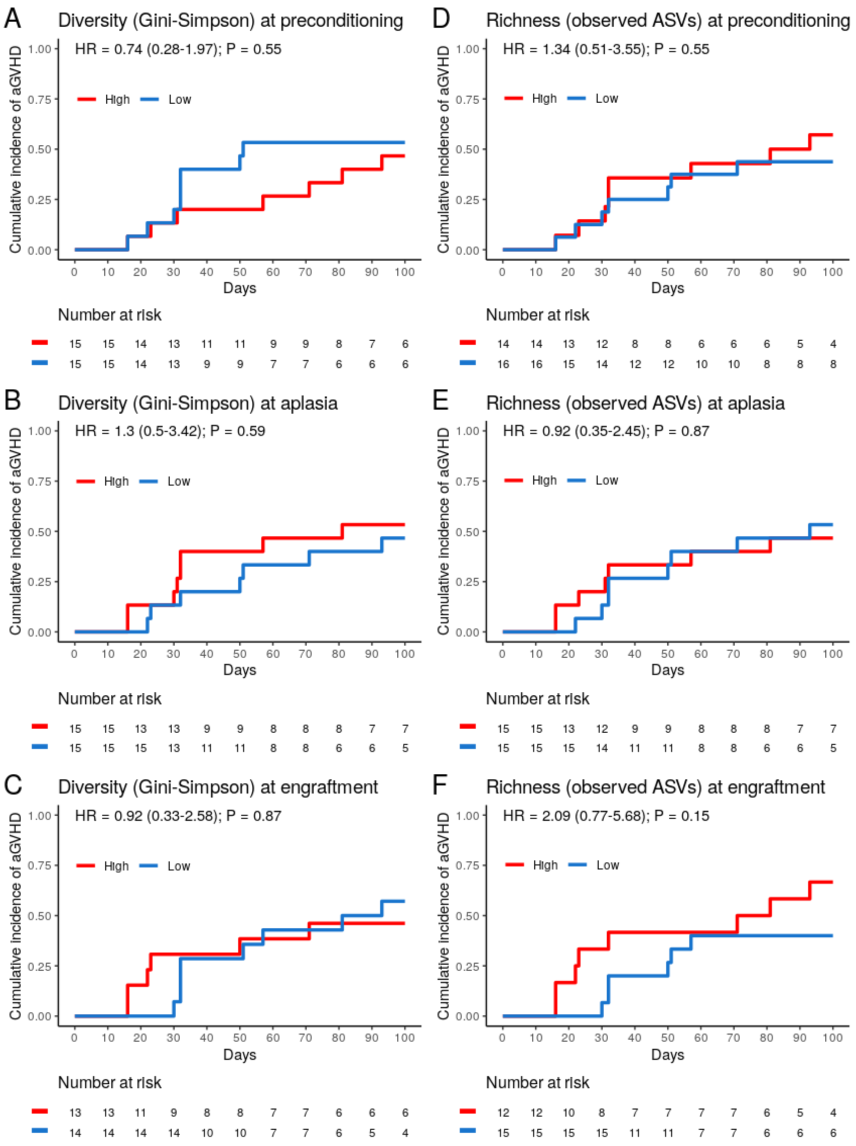


**Figure S3: Dental biofilm microbiota alpha diversity is not associated with the risk of acute graft-versus-host disease (aGVHD).** (A-C) Cumulative incidence of aGVHD with patients stratified by Gini-Simpson diversity index (High vs. Low) at preconditioning (A; n = 30), aplasia (B; n = 30) or engraftment (C; n = 27). (D-F) Cumulative incidence of aGVHD with patients stratified by the number of observed ASVs as a proxy for species richness (High vs. Low) at preconditioning (A; n = 30), aplasia (B; n = 30) or engraftment (C; n = 27).

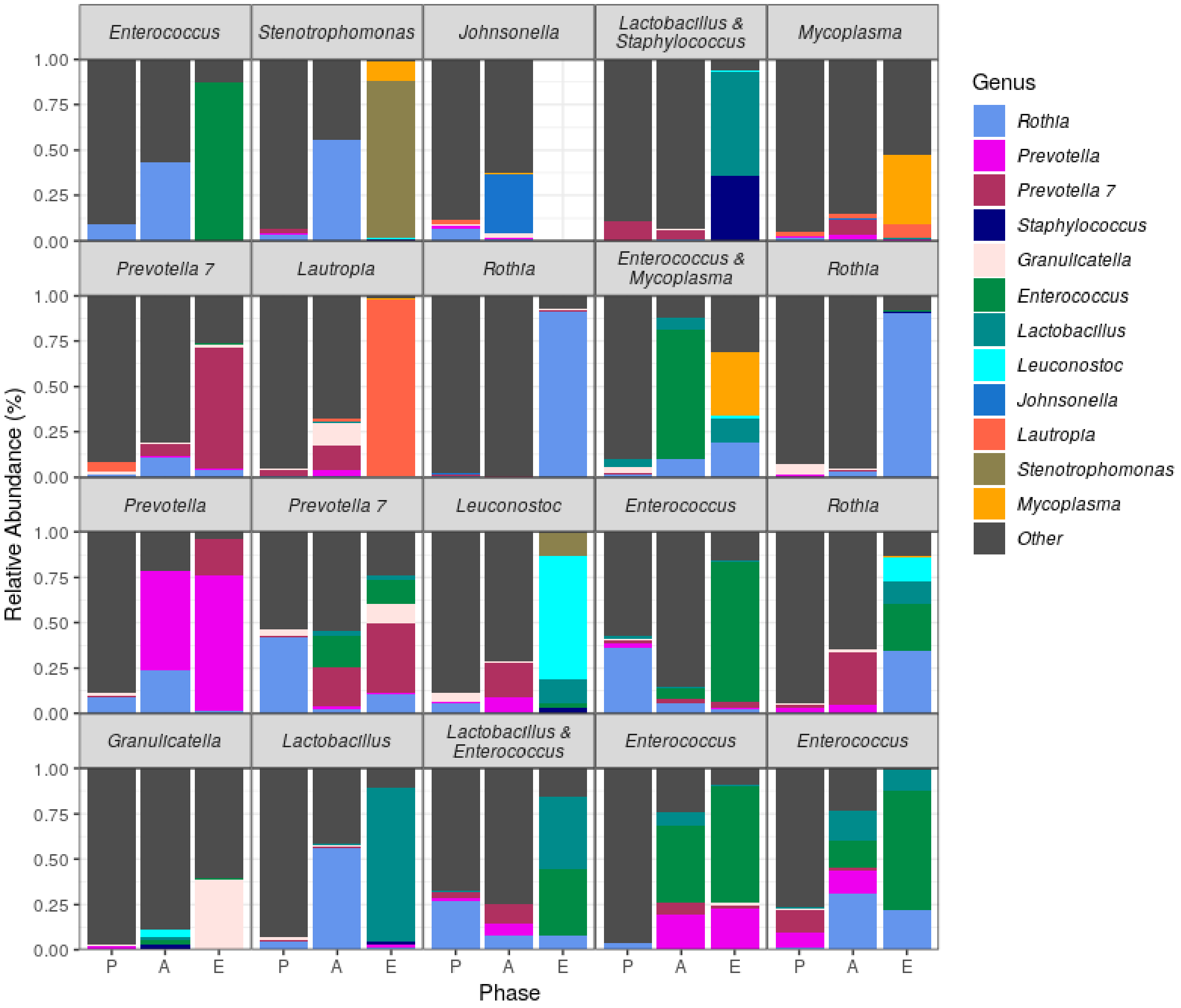


**Figure S4: Relative abundance of genera across transplantation phases for all patients experiencing genus blooming events (n = 20).** Only blooming genera are shown. Each subplot represents one patient experiencing some genus bloom, with subplot titles indicating the genera observed to bloom in such patient. P, Preconditioning; A, Aplasia; E, Engraftment.
